# Protocol for Cerebellar Stimulation for Aphasia Rehabilitation (CeSAR): A randomized, double-blind, sham-controlled trial

**DOI:** 10.1101/2024.02.05.24302365

**Authors:** Becky Lammers, Myra J. Sydnor, Sarah Cust, Ji Hyun Kim, Gayane Yenokyan, Argye E. Hillis, Rajani Sebastian

## Abstract

In this randomized, double-blind, sham-controlled trial of Cerebellar Stimulation for Aphasia Rehabilitation (CeSAR), we will determine the effectiveness of cathodal tDCS (transcranial direct current stimulation) to the right cerebellum for the treatment of chronic aphasia (>6 months post stroke). We will test the hypothesis that cerebellar tDCS in combination with an evidenced-based anomia treatment (semantic feature analysis, SFA) will be associated with greater improvement in naming untrained pictures (as measured by the change in Philadelphia Picture Naming Test), 1-week post treatment, compared to sham plus SFA. We will also evaluate the effects of cerebellar tDCS on naming trained items as well as the effects on functional communication, content, efficiency, and word-retrieval of picture description, and quality of life. Finally, we will identify imaging and linguistic biomarkers to determine the characteristics of stroke patients that benefit from cerebellar tDCS and SFA treatment. We expect to enroll 60 participants over five years. Participants will receive 15, 25-minute sessions of cerebellar tDCS (3-5 sessions per week) or sham tDCS combined with 1 hour of SFA treatment. Participants will be evaluated prior to the start of treatment, one-week post-treatment, 1-, 3-, and 6-months post treatment on primary and secondary outcome variables. The long-term aim of this study is to provide the basis for a Phase III randomized controlled trial of cerebellar tDCS vs sham with concurrent language therapy for treatment of chronic aphasia.

**Trial registration:** The trial is registered with ClinicalTrials.gov NCT05093673.

## Introduction

Aphasia is a devastating outcome and one of the leading causes of disability following stroke. Aphasia adds substantial costs to the acute [1] and chronic [2] care of individuals with stroke and is an independent predictor of subsequent functional dependence and death [3]. Anomia or difficulty with naming is the most common deficit in individuals with aphasia. Currently, the most widespread rehabilitation approach for aphasia is speech and language therapy (SALT) [4]. Although the interventions to improve naming can have benefits [5–9], a substantial number of treatment sessions is usually required to show gains, particularly in individuals with chronic large left hemisphere stroke. Therefore, to address how the treatment of aphasia might be made more effective, researchers are now using an emerging, safe, non-painful, and low-cost brain stimulation method called transcranial direct current stimulation (tDCS) [10]. There is evidence that tDCS may be useful for enhancing the effects of behavioral aphasia treatment. Evidence is growing that the add-on use of tDCS can aid in the recovery of aphasia as highlighted by international recommendations [11]. However, there is a general lack of consensus regarding the optimal electrode montage for stimulation in post-stroke aphasia. Addressing this barrier is critical for successful clinical translation.

Stimulating the residual left hemisphere region is the most common approach based on the observation that optimal recovery involves the functional re-recruitment of the remaining left-hemisphere tissue [12–16]. However, encephalomalacia filled with cerebrospinal fluid at the site of stroke affects the electrical current flow, reducing the exposure of the targeted perilesional tissue to stimulation [17]. This issue makes selection of optimal electrode locations in the left hemisphere difficult. Approaches to address this issue involve advanced electrical field modeling methods [18–20] or individualized electrode placement based on pre-treatment functional magnetic resonance imaging (fMRI) scans so that stimulation targets residual functional tissue [21–24]. However, advanced electrical field modeling and fMRI are cost-intensive and require substantial technological expertise. This would limit the incorporation of tDCS into routine speech language pathology clinical practice. We propose a novel approach to augment aphasia treatment by stimulating the right cerebellum. The right cerebellum is not only involved in cognitive and language functions (see [25–27] for reviews) but is also distant enough from typical stroke locations associated with aphasia that electrical current flow patterns are unlikely to be affected by the encephalomalacia [17]. In addition, this approach is suitable for patients who have large left hemisphere strokes and aphasia associated with bilateral hemispheric strokes.

In 2017, our group published the first study showing that cerebellar tDCS has the potential to augment aphasia treatment in a participant with bilateral middle cerebral artery infarct resulting in aphasia [28]. Subsequently, another group, utilizing a crossover study design, showed that 5 sessions of cathodal cerebellar tDCS coupled with language treatment improved verb generation immediately post-treatment in chronic post-stroke aphasia [29]. In a follow up study, we conducted a randomized, double-blind, sham controlled, within-subject crossover study in 24 chronic stroke participants with aphasia [30]. We also investigated whether there are any differences in anodal versus cathodal cerebellar tDCS on naming performance as prior studies in healthy controls have shown beneficial language effects for anodal and cathodal cerebellar stimulation [17,31–33]. Participants received 15 sessions of anodal (n=12) or cathodal (n=12) cerebellar tDCS + computerized aphasia therapy in Phase 1 followed by sham + computerized aphasia therapy in Phase 2, or the opposite order. The results of our study revealed several important findings, which have significant implications for the proposed study. First, we found that cerebellar tDCS significantly improved naming in trained (Naming 80) and untrained (Philadelphia Naming Test, PNT [34]) items immediately post-treatment, and the significant improvement in untrained naming was maintained at two months post-treatment. Second, we found that participants receiving cathodal stimulation showed significantly greater gains (compared to sham) in naming than participants receiving anodal stimulation, indicating that cathodal stimulation might be more favorable than anodal stimulation to augment aphasia treatment. Thus, these results indicate that cathodal cerebellar tDCS combined with language treatment has the potential to augment aphasia treatment.

tDCS is believed to enhance neural plasticity by temporarily modulating resting membrane potentials of neurons in targeted areas [35,36]. Anodal stimulation may lead to depolarization of the neuronal membranes resulting in greater excitability, whereas cathodal stimulation may lead to hyperpolarization resulting in lower excitability. Because the cerebellar cortex is highly convoluted and the neuronal architecture is different from cortical circuits, the polarity of cerebellar tDCS effects is not necessarily the same as the polarity of cortical tDCS effects. Animal and human studies indicate that cerebellar tDCS is most likely to produce its effects by polarizing Purkinje cells - the inhibitory output neurons of the cerebellar cortex - and thereby changing the levels/pattern of activity in the deep cerebellar output nuclei, which are the efferent targets of the Purkinje cells [37,38]. Critically, one of the deep cerebellar nuclei, the dentate nucleus, has a disynaptic excitatory connection through the thalamus to the cortical language areas. Based on this known circuitry, we hypothesize that a single session of right cathodal cerebellar stimulation will result in transient depression of Purkinje cell activity, thereby reducing the inhibitory signals that the cerebellum sends to the cortical language areas. Anodal cerebellar stimulation will exert the opposite effect, i.e., it will increase the discharge from the Purkinje cells, thereby increasing the inhibitory signals the cerebellum sends to the cortical language areas. Thus, it is plausible that multiple sessions of cathodal cerebellar tDCS will provide cortical excitation, thereby facilitating the engagement of the residual left hemisphere language areas.

In this proposal, we will combine cerebellar tDCS with semantic feature analysis (SFA) treatment for post-stroke aphasia (see [39–43] for reviews regarding SFA). SFA is a semantically-based treatment approach for naming deficits. SFA was chosen for this study for three main reasons (1) SFA has a strong potential for promoting acquisition and generalization effects for participants with anomia, (2) SFA is an effective therapy for treating naming deficits for individuals with a range of aphasia types and severities, and (3) SFA is a treatment that is frequently used by practicing speech-language pathologists (SLPs). The driving premise of SFA treatment is that when individuals generate semantic features of a target word (i.e., accessing their semantic network), they improve their ability to retrieve the target because they have strengthened access to its conceptual representation [41,44]. The theoretical mechanism by which SFA promotes generalization comes from the spreading activation theory [45] which posits that accessing/activating a particular lemma (or its features) results in activation of the lemmas of semantically related concepts. Prior studies provide strong compelling evidence that the right cerebellum, the target of our tDCS treatment, is a critical structure involved in semantic processing and naming [25–27,46–48].

Here we describe a protocol for an ongoing randomized, double-blind, sham-controlled study of cerebellar tDCS for augmenting anomia therapy in chronic aphasia. Participants are enrolled parallelly at two sites within the Johns Hopkins Rehabilitation Network: Johns Hopkins Hospital and Howard County General Hospital. We hypothesize that 15 sessions of cathodal cerebellar tDCS plus SFA will be associated with greater improvement in naming untrained pictures (as measured by the change in Philadelphia Picture Naming Test, PNT [34], 1-week post treatment, compared to sham plus SFA. For secondary outcomes, we hypothesize that cathodal cerebellar tDCS plus SFA will result in greater improvement in discourse (as measured by change in total content units (CU) and syllable per CU in picture description [49] and greater improvement in functional communication skills (as measured by change in Communication Activities of Daily Living–CADL-3 [50] compared to sham plus SFA. We also hypothesize that 15 sessions of cathodal cerebellar tDCS plus SFA will result in greater improvement on the Western Aphasia Battery-Revised (WAB-R) [51], General Health Questionnaire (GHQ)-12 [52], and Stroke and Quality of Life Scale (SAQOL-39) [53] compared to sham plus SFA.

A second aim is to identify whether neural (functional and structural) biomarkers and linguistic characteristics can predict response to cerebellar stimulation and SFA treatment. Our prior work in cerebellar tDCS in aphasia has shown that individual response to tDCS treatment is highly variable. However, little is known about how factors related to imaging and linguistic characteristics combine to induce treatment responsiveness. We will carry out resting state functional magnetic resonance imaging (rsfMRI), diffusion tensor imaging (DTI), high resolution structural imaging, and detailed linguistic testing before the start of treatment to determine whether these factors can predict response to cerebellar tDCS and/or SFA. This exploratory aim may identify stroke patients who are mostly likely to benefit from cerebellar tDCS and/or SFA. This result may have significant implications for designing a Phase III randomized controlled trial.

## Materials and Methods

### Design

This study, Cerebellar Stimulation for Aphasia Rehabilitation (CeSAR), is a Phase II trial of cathodal cerebellar tDCS plus SFA treatment vs. sham plus SFA treatment, evaluated in double-blind, randomized, sham-controlled design in chronic stroke. Participants with chronic aphasia are enrolled at two sites within the Johns Hopkins Rehabilitation Network at least 6 months after the onset of stroke. The two sites will be the Johns Hopkins Hospital and Howard County General Hospital. Sixty participants are expected to enroll over five years. The SPIRIT schedule of enrollment, interventions, and assessments is included as Fig 1. The World Health Organization Trial Registration Data Set compiled by ClinicalTrials.gov (NCT05093673) is reproduced in Table 1 (SPIRIT Item 2b). The SPIRIT checklist is included in S1 File. A full accounting of evaluations and unabridged protocol approved by the IRB is available in S2 File (September 15, 2023) and important protocol modifications will be available from the corresponding author and by viewing the ClinicalTrials.gov study entry. A sample consent form is included in S3 File.

**Figure 1.**
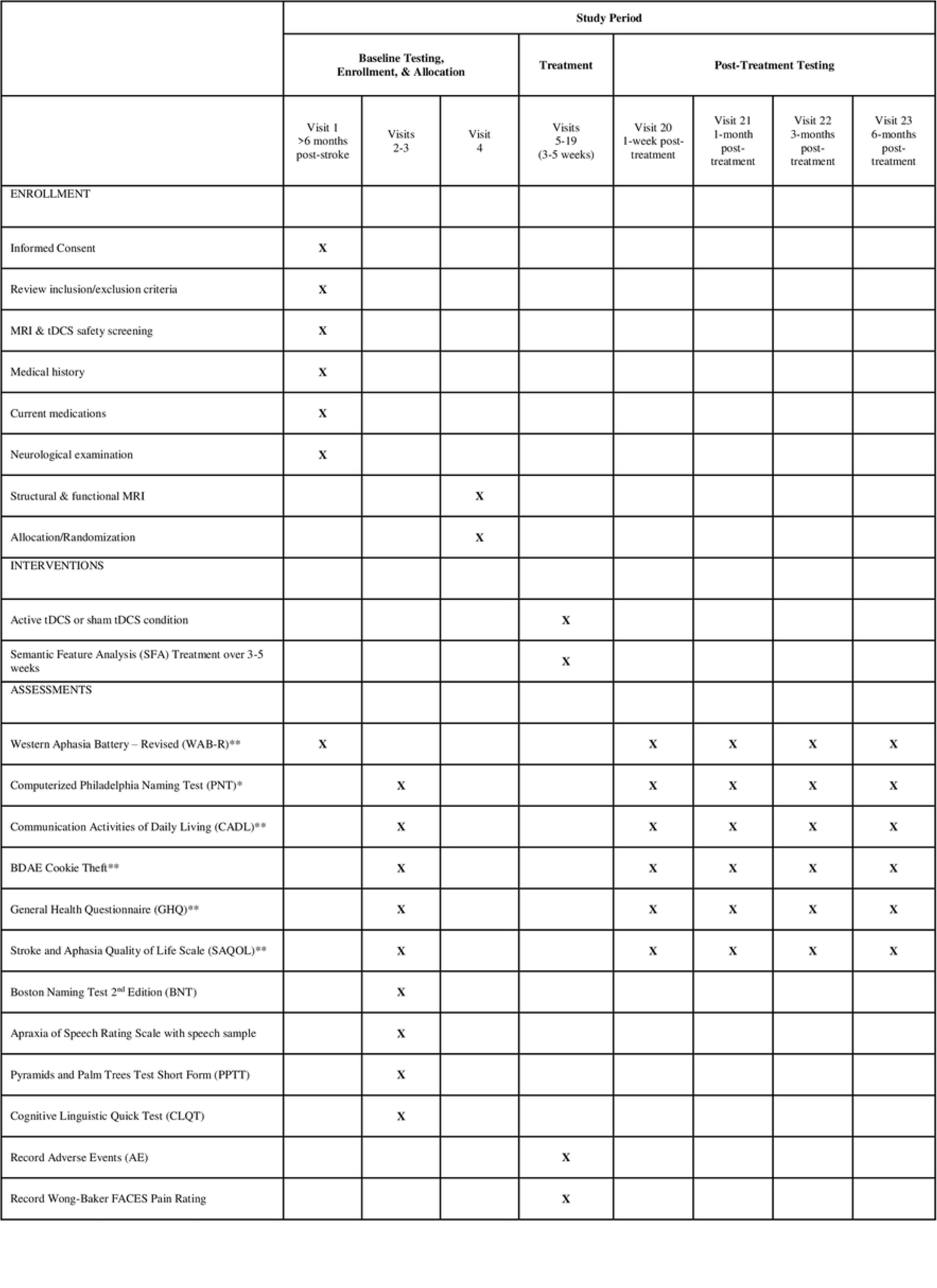
CeSAR schedule of enrollment, interventions, and assessments. *primary outcome measure **secondary outcome measures

**Table 1.**
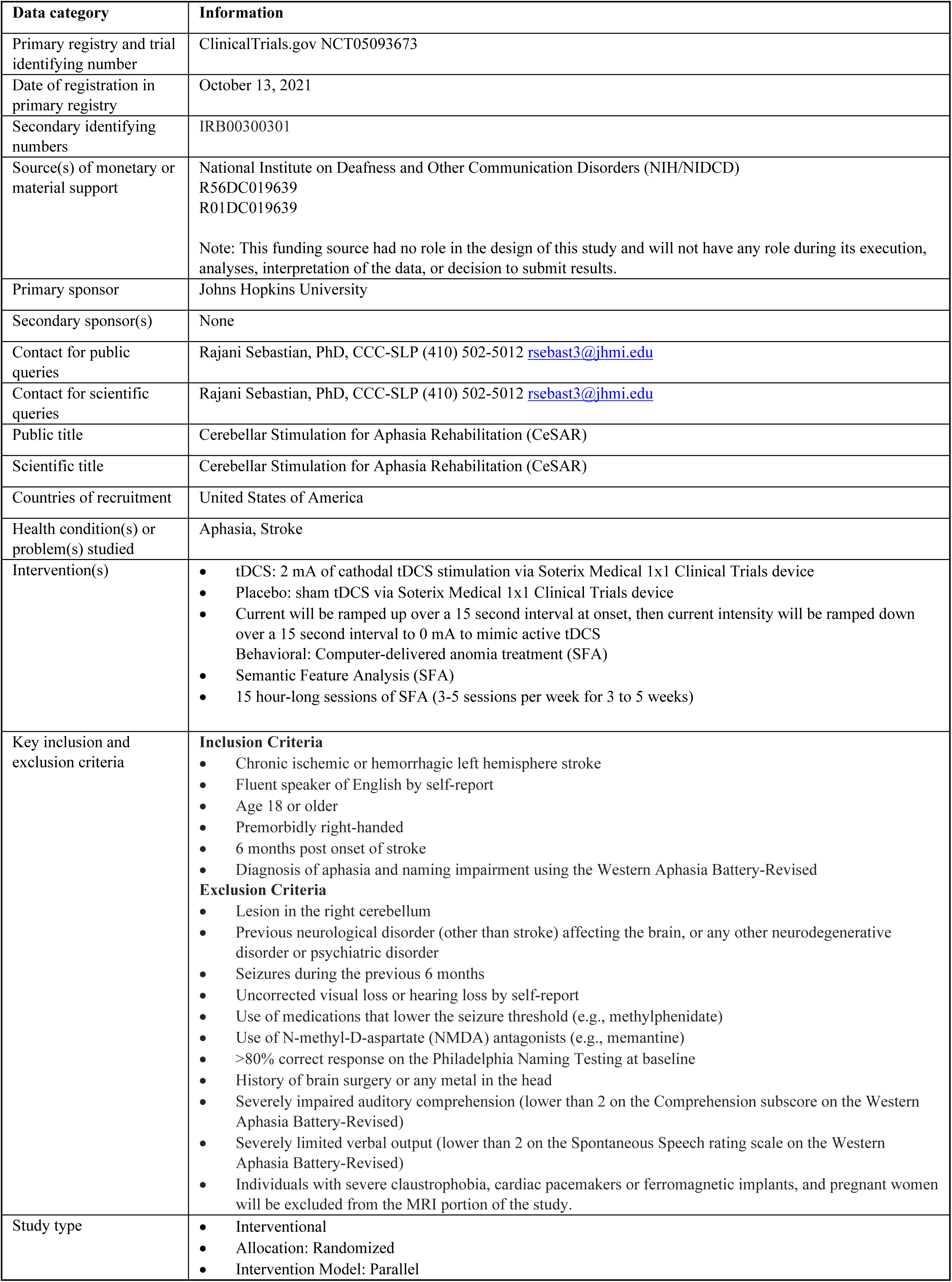
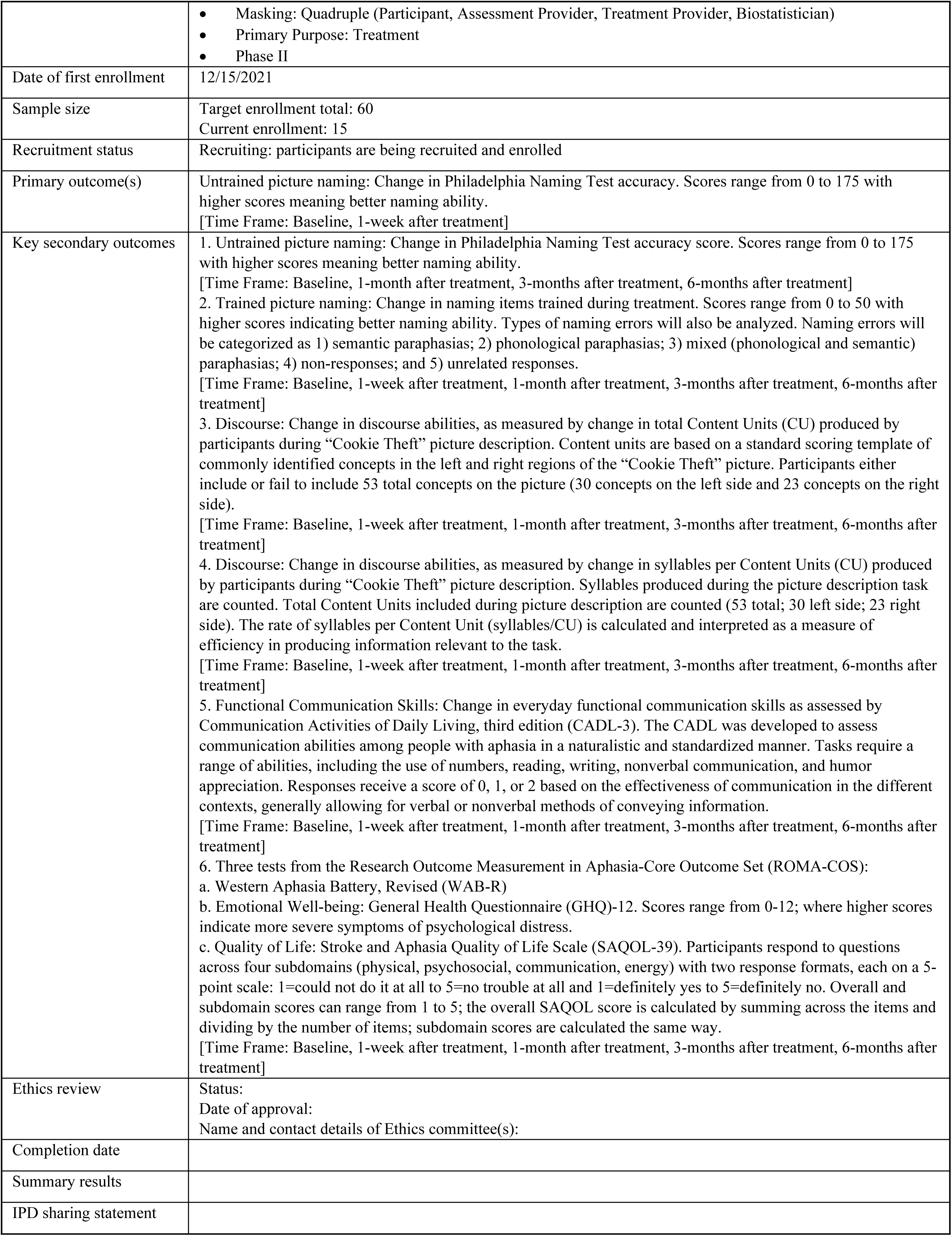
World Health Organization Trial Registration Data Set.

### Patient population-inclusion and exclusion criteria

Participants must be >6 months post ischemic or hemorrhagic left-hemisphere stroke and diagnosed with post-stroke aphasia and naming impairment using the Western Aphasia Battery-Revised (WAB-R). They must also be 18 years or older, premorbidly right-handed, and English-speaking by self-report with no lesions on the right cerebellum, with no previous neurological disorder other than stroke, or other neurodegenerative or psychiatric disorders. Individuals with seizures within the previous 6 months, those taking medications that lower the seizure threshold (e.g., methylphenidate) or N-methyl-D-aspartate (NMDA) antagonists (e.g., memantine), and those with a history of brain surgery or with any metal in the head will be excluded. We will also exclude those with uncorrected hearing or vision loss by self-report, those who score >80% on the Philadelphia Naming Test (PNT) at baseline, and those with severely impaired auditory comprehension and/or severely limited verbal output (lower than 2 on the Auditory Comprehension subscore on the WAB-R and/or lower than 2 on the Spontaneous Speech rating scale on the WAB-R, respectively). Individuals with severe claustrophobia, cardiac pacemakers or ferromagnetic implants, and pregnant women will be excluded from the MRI portion of the study.

### Inclusion Criteria

1. Chronic ischemic or hemorrhagic left hemisphere stroke
2. Fluent speaker of English by self-report
3. Age 18 or older
4. Premorbidly right-handed
5. 6 months post onset of stroke
6. Diagnosis of aphasia and naming impairment using the Western Aphasia Battery-Revised

### Exclusion Criteria

1. Lesion in the right cerebellum
2. Previous neurological disorder (other than stroke) affecting the brain, or any other neurodegenerative disorder or psychiatric disorder
3. Seizures during the previous 6 months
4. Uncorrected visual loss or hearing loss by self-report
5. Use of medications that lower the seizure threshold (e.g., methylphenidate)
6. Use of N-methyl-D-aspartate (NMDA) antagonists (e.g., memantine)
7. >80% correct response on the Philadelphia Naming Testing at baseline
8. History of brain surgery or any metal in the head
9. Severely impaired auditory comprehension (lower than 2 on the Comprehension subscore on the Western Aphasia Battery-Revised)
10. Severely limited verbal output (lower than 2 on the Spontaneous Speech rating scale on the Western Aphasia Battery-Revised)
11. Individuals with severe claustrophobia, cardiac pacemakers or ferromagnetic implants, and pregnant women will be excluded from the MRI portion of the study.

### Informed consent

A signed and dated informed consent form will be obtained from each participant. For participants who cannot consent for themselves, a legally authorized representative, such as a legal guardian or power of attorney, must sign the consent form. The consent form will describe the purposes, procedures, risks, and benefits of participation in the study, as well as the participant’s ability to withdraw consent at any time without retaliation or impact on clinical care. A copy will be given to each participant or legally authorized representative.

Once the consent form has been signed, participants will be assigned a temporary identification number for the purposes of initial screening.

All research staff authorized to obtain informed consent will have completed the Miami CITI course in the Responsible Conduct of Research and Protection of Human Subjects prior to their involvement with the study. Furthermore, they will be oriented to the study and trained by the study PI and study co-investigators who have all had extensive training and experience in the ethical and practical aspects of informed consent procedures.

### Participant confidentiality

Participation in this study should not put participants in any legal risk, even in the case of a breach of confidentiality. We will undertake every effort to keep the information in the study confidential. Participants will be assigned a code number in order to keep protected health information confidential. Consent forms and source documents will be maintained at the PI lab in a locked cabinet. All digital data will be done using participant identification numbers only and will be stored on a password-protected and encrypted format in a manner that is Johns Hopkins IRB compliant. This will include the Clinical Research Management System (CRMS), Research Electronic Data Capture (REDCap), and Johns Hopkins Microsoft One Drive. All are web-based applications designed to organize and streamline clinical research management. CRMS is integrated with Epic, Hopkins enterprise EMR, as well as Johns Hopkins IRB. This integration improves communication among study team members, stores subject enrollment information in a secure location, assists with recruitment, and allows research results to be promptly incorporated into the EMR. Everybody involved in the study will have completed the appropriate HIPAA training and are fully aware of confidentiality issues. No names will be included in any publications resulting from this work.

### Randomization

Prior to randomization, all eligible participants will receive comprehensive language and cognitive evaluations as well as MRI for those who consent and who have no contraindication. Participants will be randomly assigned 1:1 (cerebellar cathodal tDCS plus SFA treatment or sham tDCS plus SFA treatment). The randomization is stratified by study site (JHH vs Howard County), aphasia type (fluent vs. non-fluent, classified using WAB-R), and aphasia severity. Aphasia severity will be classified using WAB-R Aphasia Quotient in 4 categories (very severe aphasia: 0–25, severe aphasia: 26–50, moderate aphasia: 51–75, and mild aphasia: 76-93.8). Covariate-adaptive randomization method developed by Pocock and Simon, 1975 [54] will be implemented in REDCap. This method ensures balance on important baseline covariates by treatment arm by calculating the difference in these covariates (site, aphasia type and severity) each time a participant needs be randomized and then randomizes with high probability (80%) to the arm that corrects the imbalance on covariates.

The SLP will enter the baseline and eligibility information of a participant prior to enrollment on REDCap. If the participant’s eligibility is confirmed, then the algorithm implemented in REDCap will evaluate the treatment arm distribution in participants already randomized and then generate treatment allocation group (sham or tDCS) based on the randomization scheme. Each participant will receive a unique six-digit codes (provided by the manufacturer of the tDCS stimulator), which will instruct the stimulator to deliver either active stimulation or placebo (sham). These codes will be entered into REDCap prior to starting the study. The study coordinator will enter the codes in REDCap.

Both groups will receive semantic feature analysis treatment, a commonly used treatment for naming deficits in aphasia. It is currently unknown whether or not cerebellar tDCS augments the effect of semantic feature analysis in the chronic phase after stroke. Therefore, a sham group is justified.

### Blinding

The study is to be conducted in a double-blind manner. All participants, the members of the study team who administer the assessments, those who administer treatments, as well as the study biostatistician performing the statistical analyses will be blinded.

### Imaging

The MRI scans will be performed prior to the start of the study on a 3T Philips system at the F.M. Kirby Center at the Kennedy Krieger Institute. Imaging will be done for patients who have no MRI contraindications. Imaging will include structural and functional scans. Structural scans will include high resolution T1 and T2 weighted images, Fluid Attenuation Inversion Recovery (FLAIR) scans, and Diffusion Weighted Imaging (DWI) images. Functional scan will include resting state functional MRI.

### Treatment

Participants will receive 15 sessions of SFA treatment (3-5 sessions per week over the course of 3 to 5 weeks) and each session will be 60 minutes. Prior to the start of treatment, participants will be randomly assigned to receive either sham plus SFA or active tDCS plus SFA. The SLP will start the Semantic Feature Analysis Treatment. Participants will receive SFA treatment for 60 minutes and tDCS for the first 25 minutes. SFA treatment employed in this study will include 50 items and their relevant features from eight semantic categories. Items included in each participant’s treatment list will be determined based on performance on a picture-naming task. The naming task will consist of 200 items across eight semantic categories (food [fruits, vegetables], animals, transportation, clothing, furniture, music, sports, toys). The naming task will be administered once. To qualify for treatment, an item must be named incorrectly. To avoid effects of repeated exposure, items included on the naming task will be constrained such that they do not occur in the primary outcome variable (PNT).

Therapy tasks will be administered through a computer with clinician assistance using Microsoft Powerpoint. Participants will be trained on 7-12 items per session depending on each participant’s aphasia severity. The treatment protocol will be adapted from Doyle, Dickey and colleagues [55,56]. The treatment will proceed according to a series of steps including naming aloud the target picture, generating semantic features, naming aloud the target picture again, and generating a sentence using the target word. Participants will be asked to generate semantic features for each target picture in five categories: group [superordinate category], function [use/action], description [physical properties], context [location], and other/personal [association]. A three-level cueing hierarchy will be used to elicit features, consisting of general prompt (e.g., “How would you describe this?”), followed by a relevant directed question (e.g., “What does this feel like?”) and a binary forced-choice question (e.g., “Is this item smooth or rough?”).

tDCS will be delivered for 25 minutes using the Soterix Medical 1×1 Clinical trials device. Soterix 1×1 CT is the most advanced and customizable system for true double-blind control trials. Consistent with other studies on cerebellar tDCS [28–30,57], the current study will utilize 2 mA of cathodal tDCS stimulation generated between two 5 cm x 5 cm saline-soaked sponges. The active electrode (cathode) will be placed on the right cerebellar cortex, 1 cm under and 4 cm lateral to the inion (approximately comparable to the projection of cerebellar lobule VII onto the scalp [31]. The reference electrode (anode) will be placed over the right shoulder. For both tDCS and sham interventions, current will be ramped up over a 15 second interval at stimulation onset, eliciting a transient tingling sensation that effectively blinds the participant to treatment condition [58]. After the ramp up, in the sham condition, current intensity will be ramped down over a 15 second interval to 0 mA. Participants will rate their pain levels at the end of stimulation with the Wong-Baker FACES Pain Rating Scale (wongbakerfaces.org) [59]. In each session, participants will be asked to inform the SLP about any side effects. Participants generally tolerate tDCS well, the main reported side effects being initial tingling or itching sensations at the beginning of the session for some participants [60]. Stimulation (for both tDCS and sham conditions) will start at the same time as the aphasia treatment. Aphasia treatment will continue for another 35 minutes after the completion of 25 minutes of real tDCS or sham tDCS for a total of 60 minutes per session. Intervention for a participant will be discontinued if any of the following criteria are met: Participants will be removed from the study if they are unable to comply with task instructions or tolerate the tDCS procedure.

When the study ends, participants will continue to receive management with Dr. Argye Hillis (study neurologist) or their own neurologist as usual (generally follow-up visits approximately every 12 months). If a patient’s participation in the study ends prematurely s/he will still receive care as before. In sum, termination of the study or termination of participation in it will not affect regular therapy he or she may be receiving.

### Primary outcome

The primary outcome will be defined as the change in accuracy of naming untrained pictures measured by the Philadelphia Naming Test (PNT), one week after the end of semantic feature analysis (SFA).

### Secondary outcomes

In addition to the primary outcome, several secondary analyses will be conducted. (1) Trained Picture Naming. We will assess if tDCS has an effect on naming items trained during treatment (trained picture naming). We will assess change in trained picture naming before treatment to within 1 week after the end of treatment. Follow up testing will be done at one month, three months and six months after the completion of the treatment. In addition to assessing changes in correct naming, we will also evaluate treatment-related changes in naming errors to provide additional insight into naming recovery following cerebellar tDCS. Naming errors will be categorized as 1) semantic paraphasias; 2) phonological paraphasias; 3) mixed (phonological and semantic) paraphasias; 4) non-responses; and 5) unrelated responses. (2) Discourse. We will assess change in discourse abilities, as measured by the change in the total Content Units (CU) and syllable per CU produced by the participants during connected speech. Participants will be required to describe the Cookie Theft Picture from the Boston Diagnostic Aphasia Examination. (3) Functional Communication Skills. We will also measure changes in everyday functional communication skills assessed with the Communication Activities of Daily Living, third edition (CADL-3). (4) Finally, we will administer 3 tests from the Research Outcome Measurement in Aphasia-Core Outcome Set (ROMA-COS) at baseline, 1-week post treatment, and follow up time points. The WAB-R will be administered as a part of the baseline testing. We will also assess changes in emotional wellbeing (measured by General Health Questionnaire (GHQ)-12 and quality of life (measured by Stroke and Aphasia Quality of Life Scale (SAQOL-39).

### Data collection and quality assurance

All research staff authorized to obtain informed consent will have completed the Johns Hopkins University School of Medicine’s required training in the Responsible Conduct of Research and Protection of Human Subjects prior to their involvement with the study. Furthermore, they will be oriented to the study and trained by the study PI and study co-investigators who have all had extensive training and experience in the ethical and practical aspects of informed consent procedures.

The PI as well as the SLPs who administer baseline testing, treatments, and follow-up testing will be blinded to participant treatment assignments (described in full in the protocol provided in the Supplemental Materials). Participants will be assigned a code number in order to keep protected health information confidential. Consent forms and source documents will be maintained at the PI lab in a locked cabinet. All digital data will be done using participant identification numbers only and will be stored on a password-protected and encrypted format in a manner that is Johns Hopkins IRB compliant.

The PI (an ASHA certified SLP) will provide training to the two ASHA certified SLPs for scoring and administration of the assessment materials as well as the SFA treatment protocol. To ensure quality control, all assessment sessions and part of the treatment sessions will be videotaped. The PI will create a written protocol for clinicians regarding assessment and scoring, and to ensure consistency of delivery and adherence to SFA treatment protocol. This will reduce clinician-to-clinician variability, clinician drift, and contamination.

With respect to language assessment, the PI will be present for the first few assessment sessions to assure fidelity during assessment. This will be followed by regular monitoring to ensure adherence to assessment administration procedures. All deviations will be reviewed and clarified with the clinician to ensure that adherence is improved in subsequent sessions. Each clinician will have 20% of their total assessment sessions monitored quarterly for accurate implementation.

With respect to SFA treatment, the PI will be present for the initial few sessions to assure fidelity during treatment implementation. Following this, treatment fidelity will be monitored on a weekly basis by a member of the study team who is not providing treatment by reviewing short video-recorded segments of treatment for adherence to the SFA protocol using a Treatment Fidelity Checklist. All deviations will be reviewed and clarified with the treating clinician to ensure that adherence is improved in subsequent sessions. When session monitoring detects < 1 deviation across three consecutive samples, sessions will be monitored once bi-weekly for the remainder of the 3–5-week (3-5 sessions per week) treatment period. If session monitoring detects >1 deviations across three consecutive samples, sessions will be monitored daily until deviation is less than one. The PI and research team members meet weekly (or more often) to discuss questions about and implementation of the protocol.

To minimize the need for research-only in-person visits, telemedicine visits will be substituted for portions of clinical trial visits where determined to be appropriate and where determined by the investigator not to increase the participants risks. For the current study, we will utilize telemedicine visits when appropriate for consenting and for all the assessments visits (visits 1-3, 20-23). Prior to initiating telemedicine for study visits the study team will explain to the participant what a telemedicine visit entails and confirm that the study participant is in agreement and able to proceed with this method. Telemedicine acknowledgement will be obtained in accordance with the Guidance for Use of Telemedicine in Research. In the event telemedicine is not deemed feasible, the study visit will proceed as an in-person visit. Telemedicine visits will be conducted using HIPAA compliant method approved by the Johns Hopkins Health System and within licensing restrictions.

### Sample size estimates

Sample size was determined based on the PI’s prior crossover trial data [30]. That data was used to estimate the variability of untrained naming score. Enrolling 52 participants (26 per group) will give us 80% statistical power to detect 0.7 SD difference in change in accuracy of naming untrained items at 1-week post treatment between the study arms. This was done using Wald test for group assignment coefficient in linear regression at 0.1 level of statistical significance. The effect size (0.7SD) is a bit conservative compared to the difference observed on group comparison for 21 participants (10 in tDCS and 11 in sham) in the crossover trial data, when the tDCS was administered in Phase 1. We propose to enroll 60 participants to account for 10% attrition. However, if we have trouble meeting recruitment/retention goals, we will add Johns Hopkins Bayview Medical Center as a site.

### Statistical analyses

The primary outcome variable will be change in accuracy of naming untrained items as measured by the PNT within 1 week after semantic feature analysis ends. The analyses will follow the Intention-to-treat (ITT) principle where participants are analyzed based on the group to which they are randomized regardless of early termination, missing data or errors in randomization detected post hoc. The primary hypothesis is H_0_: mu_1_ = mu_2_ versus H_A_: mu_1_ *≠* mu_2_, where mu_1_ is the mean change in accuracy of naming untrained items between baseline and 1-week post-semantic feature analysis in the tDCS group and mu_2_ is the mean change in accuracy of naming untrained items between baseline and 1 week post semantic feature analysis in the sham group. Average Treatment Effect (ATE) will be estimated using linear regression model with change in accuracy of naming untrained items at 1 week as the dependent variable and group assignment (real tDCS versus sham) as the independent variable. ATE is estimated by the coefficient for the group assignment.

As a secondary analysis, we will consider non-parametric mixed models for analyses of functional response over time. In particular, let Y_ijk_ = u_ik_ + fk(j) + e_ij_ where Y_ij_ is the the outcome for subject i on occasion j (0, 1, 3, 6) within treatment arm k. (Thus, both i and k are necessary to identify a subject). No covariates are necessary because of the randomization. f_k_(j) is a functional model we will estimate using quadratic regression splines with knot points at each of the time points. Given there are so few time points, we will not penalize the spline fit. A non-parametric estimate of a treatment effect is given by f_2_ – f_1,_ which can show time-specific treatment effects when evaluated at specific points j. This will also demonstrate the rate (when and if) at which TCDS effects ebb. An overall effect can be estimated by simply taking the integral of f_2_ – f_1_ (i.e. the functional averaged effect over time). A null hypothesis of zero represents no time averaged effect of the treatment. Given that we will use regression splines, every estimator reduces to standard contrasts of regression parameters, and thus can be implemented in any statistical software package. Statistical analysis of secondary outcome variables will follow a similar approach as the primary outcome variable.

An additional goal of this project is to identify whether neural (functional and structural) biomarkers and linguistic characteristics can predict response to cerebellar stimulation and SFA treatment. This analysis considers moderation of treatment effects by pre-treatment baseline characteristics. The pre-treatment baseline characteristics include the following: Imaging: Structural (lesion volume, site, FA, MD), Functional (Fisher transformed connectivity values (*z* scores); Linguistic: (Aphasia Severity score as assessed by WAB-R, Naming severity score assessed by PNT). As in Hypothesis 1, we will consider both a conservative approach, using standard contrasts and median splits on the moderating variables as well as a mixed model functional approach. We will proceed in this order:

1. T-test comparing the treatment effect across median splits of the moderating variables performed separately, one at a time.
2. A non-parametric modeling approach using spline based linear models testing whether the potential moderating variables interact with the treatment effect over time. We will consider two variations of this approach:

a. One that assumes linearity
b. One that assumes non-parametric functions

### Data Monitoring body

The DSMB consists of scientists in Neurology and Public Health and will monitor safety at least semi-annually and decide if the study should continue or be terminated early. DSMB members include Kyrana Tsapkini, PhD (School of Medicine, Johns Hopkins University), John W. Krakauer, MD (School of Medicine, Johns Hopkins University), and Constantine Frangakis, PhD (Bloomberg School of Public Health, Johns Hopkins University). The study SLP in consultation with the study biostatistician will generate reports semi-annually or more frequently, as determined by the DSMB, which provide statistics on enrollment, participant status, safety data, and data quality information.

### Specification of safety parameters

The participant may stop testing or the intervention at any time. tDCS provides a non-invasive method to stimulate the cortex and cerebellum and modulate cortical/cerebellar activity via continuous, weak polarizing electrical current. This study will use the Soterix Medical 1×1 Clinical Trials system to administer tDCS. The Soterix transcranial Direct Current Stimulator Clinical Trials (1×1-CT) system is the most advanced and customizable stimulation for true double-blind control trials. It is powered by four 9-V batteries with an output of 1-2.5 milliamperes (mA). Anodal tDCS (A-tDCS) results in an increase in cortical excitability. Cathodal tDCS results in decrease in cortical excitability. To date, no serious adverse effects of tDCS have been reported in the literature as long as safety guidelines are followed [11,61]. A recent review updated and consolidated the evidence on the safety of tDCS [60]. This review shows that the use of conventional tDCS protocols in human trials (≤40 min, ≤4 mA) has not produced any reports of a serious adverse effect or irreversible injury across over 33,200 sessions and 1000 subjects with repeated sessions. This includes a wide variety of subjects, including participants with stroke. Very minor side effects such as itching, tingling, burning have been reported, as well as temporary headache, sleepiness, dizziness. However, they were generally indistinguishable from those reported by participants receiving sham stimulation. The current study will only administer 2 mA for 25 minutes per treatment session. It is important to note that tDCS does not cause significant heating effects under the electrodes, alter the blood-brain barrier, or induce edema.

Our recent study in chronic post stroke aphasia (20 min, 2mA) in 24 participants did not produce any negative effects associated with tDCS administration beyond mild itching/tingling at the beginning of the treatment session [30]. A recent large crossover trial in 36 participants with Primary Progressive Aphasia (20 min, 2mA) reported no episodes of intolerability and no serious adverse effects [62]. On the Wong-Baker FACES Pain Rating Scale, the mean pain rating for tDCS was 2.21 (standard deviation 2.48, range 0–10) and the mean rating for sham was 2.14 (standard deviation 2.13, range 0–10).

Another large, randomized control trial in 74 participants with aphasia reported 8 mild, non-serious adverse events and there were no statistically significant differences between treatment groups for number of adverse events [21]. 2 participants (6%) in the active tDCS group experienced transient scalp redness/irritation (erythema) compared with none in the sham tDCS group. On the Wong-Baker FACES Pain Rating Scale, most often individuals reported no hurt: 94% (n = 476) in the active tDCS group vs 86% (n = 511) in the sham group. The highest pain rating reported was 3 (indicating “hurts even more”), which was reported 4 times by 2 individuals (3%), both in the sham group. Taken together, all available research suggests that prolonged application should not pose a risk of brain damage when applied according to safety guidelines.

Participants may undergo MRI scanning in the present study. The effects of undergoing MR scanning have been extensively studied and there are no risks associated with an MR exam. The patient may, however, be bothered by feelings of confinement (claustrophobia), and by the noise made by the magnet during the procedure. They will be asked to wear earplugs or earphones while in the magnet.

All MRI scans will be reviewed by co-investigator and board-certified neurologist (Dr. Argye Hillis) and any suspicious abnormalities will be referred to a board-certified neuroradiologist. Please note that all of our participants, who will be recruited from the outpatient or stroke clinic, who do not have contraindication for MRI will have had a clinical MRI post-stroke. If unexpected abnormalities - incidental findings - are seen (which is unlikely, as every patient will have had a clinical MRI as part of their evaluation for stroke), the participant will be asked permission to contact the primary care physician about the abnormality, and will be offered a timely appointment with a neurologist (Dt. Argye Hillis, co-investigator) if appropriate.

Participants will be carefully screened over the phone prior to being scheduled, to assure that they meet study criteria. tDCS stimulation will be ramped up over the first 15 seconds of stimulation in order to eliminate the sensation of tingling that can occur under the electrodes during the initial moments of tDCS application. The participant may stop testing or the intervention any time. There will be emergency personnel and equipment on hand for safety.

Adverse events will be monitored during the entire visit by the study team. The families will be given telephone numbers of the study team as well. The study physician (Dr. Argye Hillis) and the DSMB will be notified immediately if any adverse events are reported. If a significant safety concern arises, participants may be unblinded in order to address it. The DSMB will determine if the adverse event is a serious adverse event. Adverse events will be monitored until they are resolved or clearly determined to be due to a subject’s stable or chronic condition or intercurrent illness. In the case of any unexpected adverse events involving risks to participants or others that are related/possibly related to the research, a Protocol Event Report will be prepared by the Study Coordinator, the PI will be informed immediately, and the IRB will be contacted within 10 days as per Johns Hopkins Medicine IRB policy; deaths will be reported within 72 hours. Also, as required by IRB policy, any unexpected adverse device effects, potential breaches of confidentiality, unresolved participant complaints will be promptly reported to the IRB. Any other adverse events that do not require prompt reporting will be summarized and reported to the IRB at the time of continuing review.

## Data Availability

No datasets were generated or analysed during the current study. All relevant data from this study will be made available upon study completion.

## Summary and concluding remarks

It is our hope that completion of this project will result in better understanding of whether and how cerebellar tDCS coupled with behavioral therapy may help individuals with post stroke aphasia. The cerebellum, which contains more than half of the brain’s neurons and a significant source of input to language as well as motor cortical regions, provides a means by which residual cortical tissue can be stimulated in stroke participants without interference from the lesion itself. However, the effect of cerebellar tDCS combined with behavioral therapy remains incompletely understood. Further, little is known about how factors related to imaging and linguistic characteristics combine to induce treatment responsiveness. We will carry out resting state functional magnetic resonance imaging (rsfMRI), diffusion tensor imaging (DTI), high resolution structural imaging, and detailed linguistic testing before the start of treatment to determine whether these factors can predict response to cerebellar tDCS and/or SFA. This exploratory aim may identify stroke patients who are mostly likely to benefit from cerebellar tDCS and/or SFA. This result may have significant implications for designing a Phase III randomized controlled trial. Trial results will be submitted to Clinicaltrials.gov no later than one year after the primary completion date. In addition, regardless of outcome, results will be disseminated in peer reviewed journals and contribute to the growing body of literature on the topic of tDCS in post-stroke aphasia rehabilitation.

## Supporting Information

**S1 File. SPIRIT checklist.**

(DOCX)

**S2 File. Protocol.**

(DOCX)

**S3 File. Consent Form.**

(PDF)

## Author Contributions

**Conceptualization:** RS, AEH

**Data curation:** NA

**Formal analysis:** NA

**Funding acquisition:** RS

**Methodology:** RS AEH GY

**Project administration:** BL, MJS, SC, JHK

**Supervision:** RS

**Writing – original draft:** BL, MJS

**Writing – review & editing:** BL, MJS, SC, JHK, GY, AEH, RS

